# Quantifying the Relative Importance of Genetics and Environment on the Comorbidity between Mental- and Cardiometabolic Disorders: A Comprehensive Analysis of National Register Data from 17 million Scandinavians

**DOI:** 10.1101/2024.02.29.24303530

**Authors:** Joeri Meijsen, Kejia Hu, Morten Dybdahl Krebs, Georgios Athanasiadis, Sarah Washbrook, Richard Zetterberg, Raquel Nogueira Avelar e Silva, John R. Shorter, Jesper R. Gådin, Jacob Bergstedt, David M. Howard, Weimin Ye, The iPSYCH Consortium, Yi Lu, Unnur A. Valdimarsdóttir, Andrés Ingason, Dorte Helenius Mikkelsen, Oleguer Plana-Ripoll, John J. McGrath, Nadia Micali, Ole A. Andreassen, Thomas M. Werge, Fang Fang, Alfonso Buil

## Abstract

Mental disorders (MDs) are leading causes of disability and premature death worldwide, partly due to high comorbidity with cardiometabolic disorders (CMDs). Reasons for this comorbidity are still poorly understood. We leverage nation-wide health records and complete genealogies of Denmark and Sweden (n=17 million) to reveal the genetic and environmental contributions underlying the observed comorbidity between six MDs and 14 CMDs. Genetic factors contributed about 50% to the comorbidity of schizophrenia, affective disorders, and autism spectrum disorder with CMDs, whereas the comorbidity of attention-deficit/hyperactivity disorder and anorexia with CMDs was mainly or fully driven by environmental factors. These findings provide causal insight to guide clinical and scientific initiatives directed at achieving mechanistic understanding as well as preventing and alleviating the consequences of these disorders.

---

Individuals with mental disorders (MD) present substantially higher rates of various medical conditions^1^ and mortality^2^ as compared to the general population. Indeed, individuals with a mental disorder diagnosis face a 7- to 10-year shorter life expectancy^3^. This is mainly due to the 2- to 3-fold increased risk of premature death from somatic comorbidities such as cardiovascular and cardiometabolic disorders (CMD)^3–5^, including coronary artery disease (CAD)^6^, stroke^7^, and heart failure (HF)^8^. These disorders are often preceded by other related conditions such as type-2 diabetes (T2D), high cholesterol, obesity, and hypertension. Later epidemiological studies^1,9,10^ systematically leveraging population-wide, healthcare registers confirmed and extended the positive MD-CVD association and found it to be bidirectional, i.e. also mentally healthy individual at the time of CVD onset, go on to develop mental illness more frequently than expected by chance.

Importantly, risk increase may vary considerably across pairs of individual mental and cardiovascular diagnoses. In fact, individuals diagnosed with anorexia nervosa (AN), attention-deficit/hyperactivity disorder (ADHD), or an affective disorder (AFF) are more likely to be diagnosed with atrial fibrillation later in life than individuals with schizophrenia (SCZ) or autism spectrum disorder (ASD), whose risk may be lower than the background population^1^. The observations of complex patterns of comorbidities indicate considerable diversity in the underlying causes, which may include both shared genetic architectures and environmental exposures such as prescription drugs, substance abuse, socioeconomic conditions, and traumatic live events.

An important step toward designing effective prevention and treatment strategies to combat these comorbid conditions would be to effectively discriminate heritable genetic effects and environmental risk exposures for each comorbid pair of MD and CMD. When the comorbidity is driven mainly by environmental factors, clinical approaches will be limited to interventions in the environment. However, when genetics plays an important role, therapeutic approaches could also benefit from precision medicine tools such as genetic risk prediction models.

Notably, progress is complicated by extensive correlation among, as well as between, environmental risk exposures and inherited disease liability, challenging the identification of causative factors^11^. Previous studies have found a substantial genetic overlap between CMD and MD^12–15^, which suggests genetics contribute to the MD-CMD comorbidity. However, these predominantly molecular genetic studies are affected by extreme sampling of cases suffering from comorbidities other than the index diagnosis under study, and of control participants recruited among overly healthy subjects, and thus, they are genuinely unrepresentative of the background population^16^. These previous studies report genetic correlations as a measure of shared genetic effects between two disorders but do not investigate the relative importance of genetic and environmental factors on the comorbidity between disorders.

Nationwide genealogies cross-referenced with sociodemographic and comprehensive healthcare registers from an exclusively public healthcare system do provide an attractive possibility to perform large-scale and population-true analyses allowing to discriminate shared heritable from environmental factors from across individuals in extended families. Here we leverage nationwide healthcare registers and population-complete genealogies spanning 4 generations of Denmark and Sweden for 17 million individuals to quantify the genetic contribution underlying the comorbidity between 6 mental disorders and 14 cardiometabolic disorders. For each country, we first estimate the incidence of 6 mental disorders and 14 cardiometabolic diseases in the general population and for individuals with affected relatives. Next, we determine the heritability for each single disorder as well as the genetic correlations between all pairs of diagnoses. Finally, we quantify the relative contribution of genetic factors for each of the comorbid MD and CMD constellations.

## Results

Disentangling and quantifying the causes of clinically important and etiologically complex constellations of diseases, such as comorbid mental disorders with cardiovascular diseases, relies on the ability to reliably observe manifestations of both classes of diseases in large populations of related individuals over decades. However, differences in age-at-onset, length-of-observation periods, population structure, healthcare provision and ascertainment biases have traditionally compromised our understanding of the classical nature-nurture duality of causalities.

Here we circumvent these challenges by leveraging two unique, near-complete population genealogies: those of Denmark and Sweden, including 6 and 11 million individuals across 4 generations, respectively. The two populations have been served for decades by similar, public healthcare systems. Clinical diagnoses of diseases have been systematically and uniformly recorded for 39 and 43 years, and data made available for analyses alongside population-wide information of individual-level, familial relatedness.

To estimate heritability, genetic correlations, and environmental risk components across the six selected mental disorders and 14 mostly common and severe cardiovascular disorders, we first determined the cumulative incidence function (CIF) across birth years^17,18^. As exemplified for ADHD (Figure 1A) and for all six mental disorders (Table S1), CIFs vary systematically across birth-years, and year-based CIFs are therefore included in all subsequent analyses of heritability and genetic correlations as outlined in detail below.

**Figure 1:**
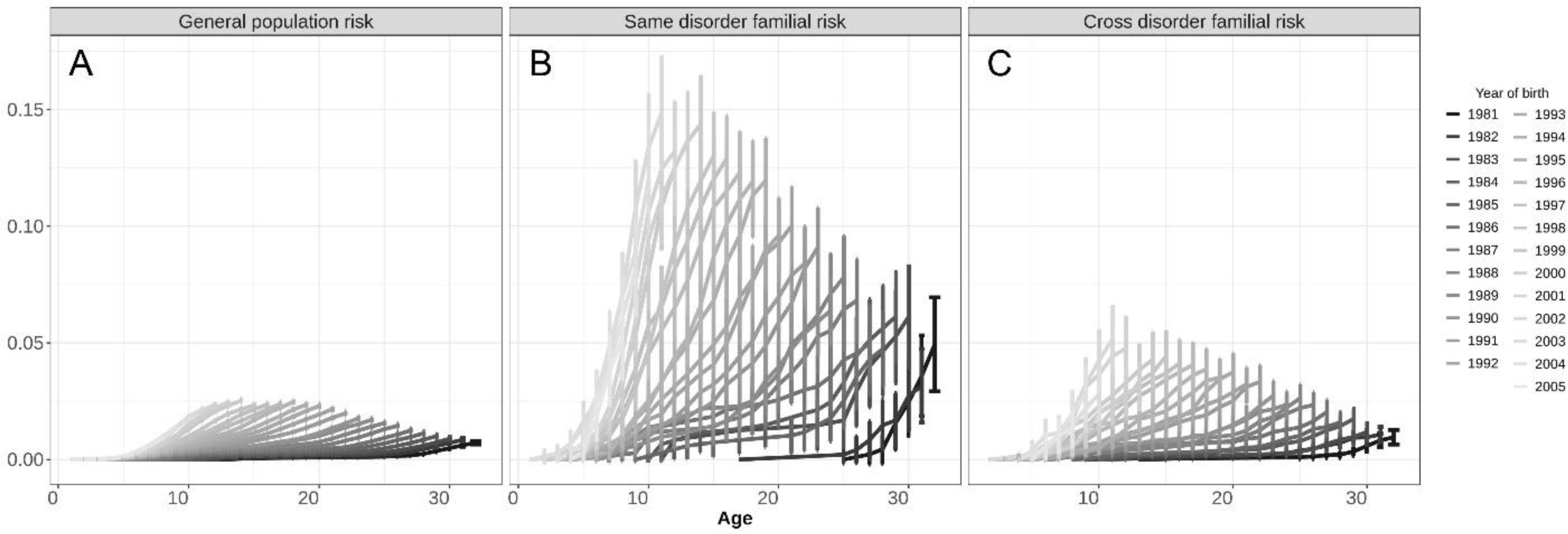
Cumulative incidences and 95% confidence intervals of ADHD for individuals born 1981-2005 using medical records up to 2012 stratified by year of birth. *Cumulative incidences are shown for: A.) the general population, B.) individuals with at least one full-sibling diagnosed with ADHD, C.) individuals with at least one parent diagnosed with type-2 diabetes*.

Notably, the CIF for individuals with a sibling with ADHD (Figure 1B), as well as the CIF for individuals with a parent diagnosed with Type-2 diabetes (Figure 1C) are markedly increased relative to the CIF of ADHD for the entire population (Figure 1A). These observations are consistent with ADHD being highly heritable and with significant genetic-correlation between ADHD and type-2 diabetes^19^. All Danish CIFs are listed in Table S1.

We applied this analytical approach to systematically estimate heritability and genetic correlations for all six mental and 14 cardiovascular- and metabolic disorders in the Danish and Swedish populations. As shown in Figure 2, heritability estimates range from 25% to 75% and were strongly correlated between the two countries (Figure S1A, Table S2) and are broadly concordant with existing studies (Table S3). Similarly, we estimated genetic correlations for each of the 84 diagnostic MD-CMD pairs in both populations and observed 28 significant correlations (Bonferroni *p*<5.98x10^-4^) after meta-analyzing the highly similar country-specific estimates (Figure 3 and Figure S1B). With the notable exception of AN, the genetic correlations of the mental disorders with the 14 cardiometabolic disorders were positive. They were larger for ASDs and bipolar disorder (BD) than for ADHD and AFFs (Table S4).

**Figure 2:**
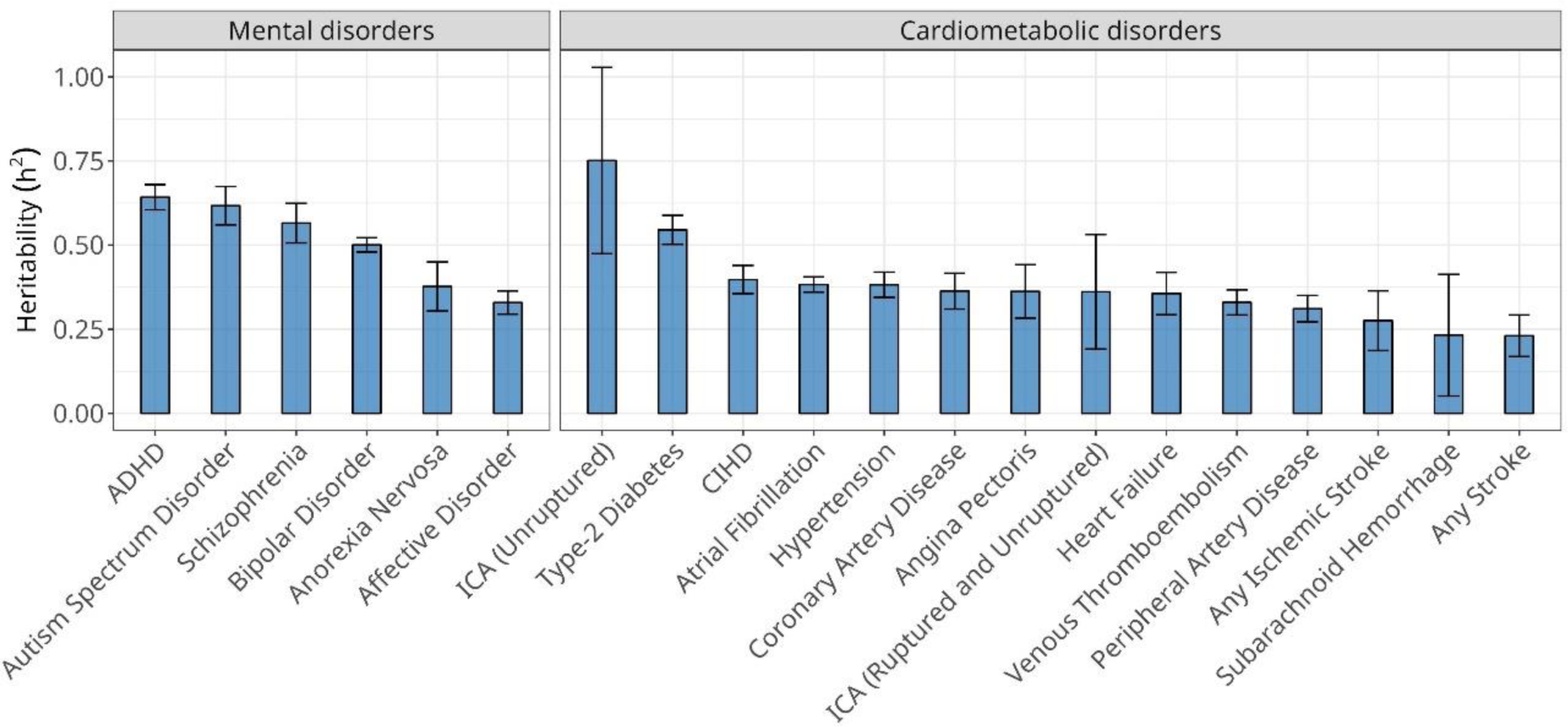
Meta-analysis of Danish and Swedish narrow sense heritability (h^2^) estimates and 95% confidence intervals of mental- and cardiometabolic disorders. *ADHD = attention deficit/hyperactivity disorder, ICA = intracranial aneurysm, CIHD = chronic ischemic heart disease*

**Figure 3:**
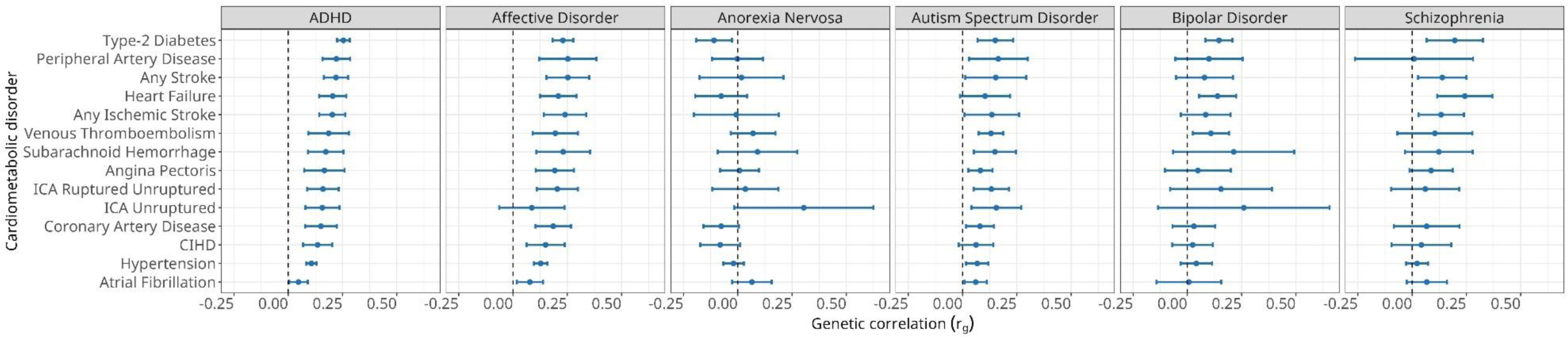
Meta-analysis of Danish and Swedish genetic correlations (rg) estimates between mental- and cardiometabolic disorders and 95% confidence intervals calculated using national register data. *ADHD = attention deficit/hyperactivity disorder, ICA = intracranial aneurysm, CIHD = chronic ischemic heart disease*

Genetic correlation, while estimating the degree of overlap of the genetic architectures of two disorders, may account for only a neglectable fraction of the observed comorbidity. This scenario may occur for disorders with high genetic correlations when the heritability of one or both disorders is modest, as shown in simulated data in Figure S2. To determine the relative contributions of heritable genetic variants and environmental risk factors to MD-CVD comorbidities, we applied quantitative genetic methods that combine the estimates of heritability and genetic correlation for each pair of disorders. As a measures of comorbidity, we used hazard ratios previously determined in the Danish population^1^, allowing us to estimate the relative heritable and environmental components for 35 of the 84 pairs of MD-CVD. As shown in Figure 4, the environmental component is comparable and, in most cases, markedly higher than the genetic component of the comorbidity. Notably, the genetic components for the comorbidity between all CMDs with AN was negligible and it was also very low with ASD. For all MDs, the comorbidity with hypertension and atrial fibrillation has a lower genetic components than for other CMDs. As observed for ASD, negative phenotype correlation with CMDs is driven by protective environmental factors. All estimates (i.e., hazard ratios, cardiometabolic prevalence’s, rp, G, E) including SE, z-scores, CIs, and p-values are reported in Table S5.

**Figure 4:**
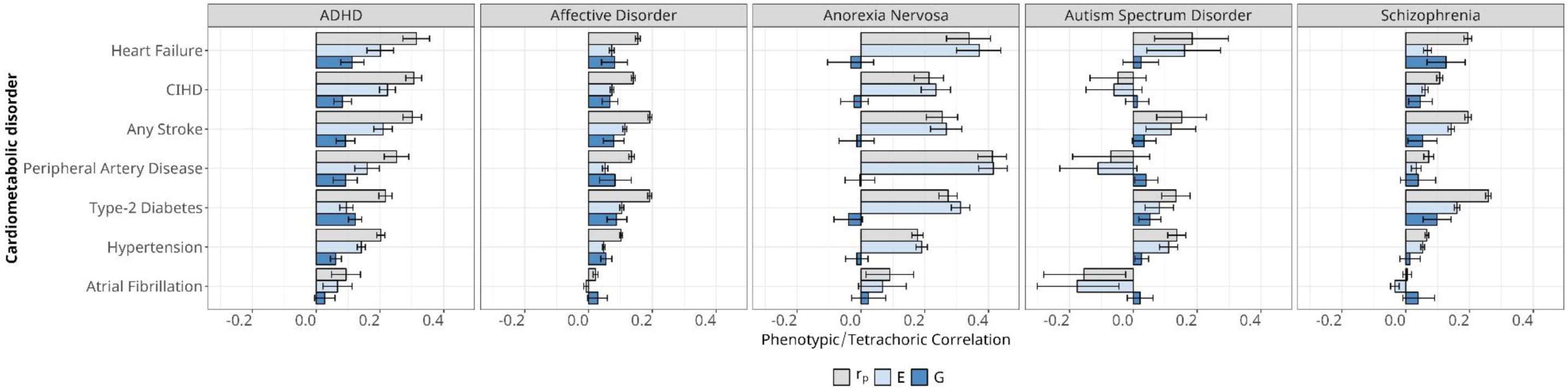
Quantification of the contribution of genetic (G) and the non-genetic (E) factors to the observed phenotypic correlation **(rp)** between mental- and cardiometabolic disorders using register based genetic correlations (rg) and heritability (h^2^) estimates. ***Estimates of rp were selected from Momen et al.^1^*.**

In summary, the observed MD-CMD comorbidity patterns are mostly explained by shared environmental rather than heritable factors, a trend that is particularly pronounced for AN and ASD, as well as for hypertension and atrial fibrillation.

### Comparable estimations of heritability and genetic correlation using genotype information

Since most studies on heritability and genetic correlations are performed using GWAS summary statistics, we compared our results with those obtained using SNP-based heritabilities (h^2^) and genetic correlations (rg SNP). We used LDSC and summary statistics from well-powered GWAS (Table S6). To make results comparable with our register-based results we used MD GWAS for the large Danish cohort iPSYCH 2012^20,21^. We converted these estimates to the liability scale^22^.

On a liability scale, the median of h^2^ across all MD combined was 0.17 (SD=0.03), lowest for BD (0.13; 95% CI 0.03-0.24) and highest for ADHD (0.23; 95% CI 0.2-0.27) for ADHD. The h^2^ across CMD was lower with a median of 0.09 (SD=0.06), lowest for heart failure (2.3x10^-2^; 95% CI=1.93x10^-2^-27x10^-2^) and highest for unruptured ICA (0.22; 95% CI 0.13-0.32). Thus, relative to the register-based heritability estimates (h^2^), the h^2^ were lower for MD (63%) and CMD (72%). Correspondingly, h^2^ for the individual MD and CMD disorders), were significantly smaller than the register-based h^2^ estimates, except for SAH and ICA (Figure S3 and Table S7).

Regarding SNP-based genetic correlations, we observed 21 substantial and significant genetic correlations, of which 12 were associated with ADHD, 6 with AFFs, and the remaining three with AN, ASD, and SCZ (Bonferroni multiple testing correction threshold *p<*5.95x10^-4^). These SNP-based genetic correlations were not significantly different from the register-based Danish estimates (Figure S4, Bonferroni *p<*5.95x10^-4^), nor from Danish and Swedish meta-analysed genetic correlations (Table S8). To test if these results are dependent on the specific GWAS used, we repeated the analyses using the larger PGC meta-analysis summary statistics and observed no significant differences compared to the use of the Danish-only GWAS summary statistics (Figure S5).

The high concordance between the register- and the SNP-based estimates confirms a substantial shared genetic contribution, based on common genetic variants between many MDs and CMDs. However, since the genetic contribution to comorbidity also depends on the heritability of both diseases, the comorbidity partition using SNP-based estimates largely underestimates the genetic component (Figure S6). This fact highlights the importance of family-based estimates derived from large national registers.

## Discussion

One of the main challenges in healthcare is to understand the causes of complex patterns of comorbidity. Here we provide the first systematic and comprehensive analyses to discriminate and quantify environmental and heritable causes underlying comorbidity between the clinically important disease domains of mental and cardiometabolic disorders. Leveraging unique genealogies and cross-referenced, nationwide healthcare registers holding near-complete records from the public healthcare systems in Denmark and Sweden, we document that the notable comorbid manifestations of single pairs of mental and cardiometabolic disorders are more environmental (50-70%) than heritable in origin. Importantly, these findings show that the nature-nurture origin of comorbid presentations cannot be deduced from estimates of the genetic correlation of the corresponding disorders. Moreover, we show that the relative contribution of genetic and environmental causes to the observed comorbidities between MD and CMD varies largely and is specific for each disease pair.

We analysed six mental disorders and 14 cardiometabolic diseases, and we observed large variability between the relative contribution of genetics and environment to the comorbidity of the 84 disease constellations. We observed that the observed comorbidity between anorexia nervosa and most CMDs can be explained exclusively by non-genetic effects, while the genetic component plays a substantial role (around 50%) in the comorbid presentations of affective disorder with most of the examined CMDs. Constellations involving CMDs with ADHD or schizophrenia show intermediate genetic contribution of around 30%. In summary, the environmental and genetic causes of comorbidity vary considerably across pairs of disorders and cannot be deduced simply from the heritability of the individual disorders (or their genetic correlation); rather insight into the origin of comorbidity requires individual analyses and interpretation. This general principle is illustrated by two pairs of diagnoses, that of AFF and hypertension, and between ASD and T2D. Both pairs are characterized by a low genetic correlation of 0.1; however, in the first pair, 50% of the comorbidity can be explained by genetic factors, whereas in the second pair the genetic contribution is not different from zero. This shows that the same genetic correlation estimate could result in different estimates for the relative importance of genetics and environment on the comorbidity between two disorders. Therefore, to appreciate both the quantitative and qualitative composite nature of the causality underlying comorbid presentations, each disease-pair should be considered individually.

The interpretation of these results requires some discussion on the nature of environmental effects. There are two diverse types of environments, one related to the behaviour of the individual itself while the other is mediated by external agents. The environments created by the individual behaviour may stem from the mental disorders themselves, e.g., physical immobility, irregular sleep, and overeating are characteristic of many MDs, while compulsive behaviour and derived alcohol misuse, smoking, and overeating are specifically linked to ADHD. External sources of environmental risk in mental disorders include second-generation antipsychotics, most notably clozapine, prescribed to individuals with SCZ. These medications provoke overeating, malnutrition, obesity, long-QT, dyslipidaemia, and electrolyte deregulation, all of which are well documented risk factors for CMD^23^. As the most extreme example, we provide evidence that the comorbidity between AN and T2D can be explained exclusively by environmental factors. In this case, the combination of an individual’s behaviour and external environmental factors associated with the treatment for AN could be responsible for the AN-T2D comorbidity. For example, dietary restriction and malnutrition in individuals with AN causes long-term effects on endothelial tissues, increasing the risk of T2D^24^. Furthermore, individuals with active AN are subject to a metabolic system that is no longer working at normal capacity, which increases the risk for T2D when AN is being treated using a high-calorie diet during inpatient refeeding. An additional environmental challenge in individuals with AN is smoking, which has been shown to be more prevalent due to its appetite suppressant qualities^25^.

Our results offer a much-needed discussion of the mechanisms of comorbidities and suggest further lines of investigation into the causes of MD-CMD comorbidity. The observation of a substantial genetic component in some MD-CMD comorbidity constellations, justifies attempts to identify the genetic determinants of comorbidity between specific MD-CMD pairs that may have clinical implications. First, studies aiming to discriminate genetic underpinnings of single disorders from the genetic determinants of comorbid conditions would inform us about pleiotropic disease processes across different organ systems, as well as guide drug discovery or repurposing. Our results also highlight the importance of environmental factors as major causes for comorbidity between many MD-CMD pairs. However, new studies are needed to identify the relevant environmental exposures for each MD-CMD pair.

A strength of this study is the use of nationwide registers in Denmark and Sweden. These countries share many similarities in history, (genetic) ancestry, cultural characteristics, and their government funded healthcare system. However, clinical variation such as different recoding of diagnoses periods, use of different ICD versions^26^, and varying diagnostic practices have resulted in country-specific time trends. Despite this, replication analysis of heritability and genetic correlation estimates derived using the larger Swedish registers showed no significant difference from the Danish estimates. The high transferability between the two genealogies adds validity to the findings on heritability and genetic correlations. To complement our register analysis, we computed SNP-based estimates of heritabilities and genetic correlations leveraging molecular genetic data on the Danish population-based sample (iPSYCH 2012 cohort). To our knowledge this is the first comparison of heritabilities and genetic correlations based on genotypes and register data carried out in the same population.

Our study has also some limitations. First, the study leverages secondary care hospital diagnoses from 1968 onwards, and due to truncation, some diagnoses introduced more recently are underrepresented in the older individuals. However, our strategy of estimating heritability and genetic correlations by by-year meta-analyses partially controls for these biases in the data. Second, we did not quantify the influence of shared environment in close relatives on the heritability and genetic correlation estimates, and therefore these estimates might be slightly inflated, although previous studies suggest that such effects are small^27^. Third, in our model we assume no interaction between the genetic and non-genetic factors that contribute to the comorbidity between mental and cardiometabolic disorders, which may affect the estimated effect sizes. Finally, while the Danish sample size is large, and results replicated well in the Swedish register data, it is uncertain whether our results translate to non-Scandinavian countries and populations using different healthcare systems inside and outside Europe. However, we think it is reasonable to expect that countries with similar universal health systems and levels of development would give similar results^26^.

To the best of our knowledge this study is the first to quantify the relative contribution of genetic and environmental factors to the comorbidity of different constellations of mental and cardiometabolic diseases by integrating health registers and genealogical information from two countries and cross-referencing with molecular genetic data from a representative population sample. This work provides a foundation to guide future precision medicine by helping to implement clinical interventions to prevent and treat CMD in individuals with mental disorders.

## Ethical approval

The use of Danish data was approved by the Danish Health Data Authority (project no. FSEID-00003339) and the Danish Data Protection Agency. By Danish law, informed consent is not required for register-based studies. The use of Swedish data was approved by the regional ethics review board in Stockholm, Sweden with DNR 2012/1814-31/4.

## Data sharing statement

This work is based on Danish register data that are not publicly available due to privacy protection, including General Data Protection Regulation (GDPR). Only Danish research environments are granted authorization. Foreign researchers can, however, get access to data. Further information on data access can be found at https://www.dst.dk/en/TilSalg/Forskningsservice or by contacting the senior corresponding authors. Data from Swedish registers are not available for sharing due to policies and regulations in Sweden. Swedish register data are available to all researchers through applications at Statistics Sweden (SCB, https://www.scb.se/en/) and The National Board of Health and Welfare (Socialstyrelsen, https://www.socialstyrelsen.se/)

## Acknowledgement

This research was supported by: the European Union’s Horizon 2020 Research and Innovation Programme: the “predicting comorbid cardiovascular disease in individuals with mental disorder by decoding disease mechanisms” project (CoMorMent, grant number 847776, to JM, JB, AB, UAV, DHM, YL, TMW, OAA and FF); the Danish National Research Foundation (grant number DNRF148); the US National Institutes of Health study on extreme MDD (R01 MH123724, to JM, JRS, YL, and AB); the Sir Henry Wellcome Postdoctoral Fellowship (Reference 213674/Z/18/Z, to DMH); the Research Council of Norway (RCN grants 324499, 324252, 223273, to OAA); the Stiftelsen Kristian Gerhard Jebsen (grants SKGJ-MED-008 and SKGJ-MED-021, to OAA); the Laureate Grant Award from the Novo Nordisk Foundation (Grant No: NNF22OC0071010, to NM); European Research Council Consolidator grant (StressGene, Grant nr. 726413 to UAV), and Icelandic Research fund (to UAV). The iPSYCH Initiative is funded by the Lundbeck Foundation (Grant Nos. R268-2016-3925, R102-A9118, and R155-2014-1724), the Mental Health Services Capital Region of Denmark, University of Copenhagen, Aarhus University, and the University Hospital in Aarhus. Genotyping of iPSYCH samples was supported by grants from the Lundbeck Foundation, the Stanley Foundation, the Simons Foundation (Grant No. SFARI 311789), and National Institutes of Mental Health (Grant No. 5U01MH094432-02). The iPSYCH Initiative uses the Danish National Biobank resource that is supported by the Novo Nordisk Foundation. We like to thank the Psychiatric Genomics Consortium working groups (major depressive disorder, attention-deficit/hyperactivity disorder, autism spectrum disorder, schizophrenia, bipolar disorder, and eating disorders) for contributing genome-wide association summary statistics data. Finally, we like the thank Hakon Heimer for and editing the final version of the manuscript.

## Declaration of interests

NM receives an honorarium to serve as associate editor on the European Eating Disorders Review board. OAA is a consultant to Corteechs.ai and received speaker’s honorarium from Janssen, Sunovion and Lundbeck. UAV declares receiving support from EPA2023, ISTSS2022 as keynote speaker, and serves on a NordForsk expert committee on Long COVID.

## Online methods

### Integrative Psychiatric Research Consortium 2012 (iPSYCH2012) cohort

The iPSYCH2012 cohort is a well-documented extensively cited cohort^20^. In short, individuals born between 1981 and 2005 (n=1,472,762) were considered for ascertainment, representing the entire population of Denmark born in that timeframe. Of these 30,000 were randomly sampled, regardless of (psychiatric) disorder status, to create an unbiased population representative control group. Using information ascertained from the Danish Civil^28,29^, National Patient^30^ and/or Psychiatric Central Research Registers^31^ 57,764 design cases were selected with indications of clinical diagnoses of mental health disorders. In total, 87,764 individuals were selected to form the cohort. Indications are based on International Classification of Disease (ICD) codes representing the clinical diagnosis associated with an instance of care provided at one of many psychiatric facilities throughout Denmark. The following six case groups as having at least one indication with the corresponding ICD10^32^ (or equivalent ICD8^33^) codes were defined: attention deficit hyperactivity disorder (ADHD: F90.0), anorexia nervosa (AN: F50.0, F50.1), autism spectrum disorder (ASD: F84.0,F84.1, F84.5, F84.8, F84.9), affective disorder (AFF: F30-F39), bipolar disorder (BD: F30-31), and schizophrenia (SCZ: F20). Of all selected individuals a dried neonatal heel prick blood spot was obtained from the Danish Neonatal Screening Biobank^34^. Individuals were removed when no blood spot could be obtained. The use of this data is according to the guidelines provided by the Danish Scientific Ethics Committee, the Danish Health Data Authority, the Danish data protection agency, and the Danish Neonatal Screening Biobank Steering Committee. For each dried bloodspot the DNA was extracted and amplified followed by genotyping using the Infinium PsychChip v1.0 array^20^. Of 9,714 bloodspots not DNA could successfully be genotyped and therefore the individuals were excluded from the study. A subset of good quality SNPs were phased into haplotypes using SHAPEIT3^35^ and imputed using Impute2^36^ with reference haplotypes from the 1,000 genomes project phase 3^37^. Genotypes were checked for imputation quality (INFO >0.2), Hardy-Weinberg equilibrium (HWE; p<1x10^-6^), association with genotyping wave (p<5 × 10^−8^), association with imputation batch (p<5×10^−8^), differing imputation quality between subjects with and without psychiatric diagnoses (p<1×10^−6^), and minor allele frequency (MAF>0.01). Finally, we extracted unrelated individuals of European ancestry leaving 77,082 individuals.

### Danish nationwide registers

The *Danish Civil Registration System* was established in 1986 and contains detailed information pertaining to sex, date of birth, parental links, and continuously updated information on vital status (e.g. migration or death) for all individuals alive and living in Denmark for the past seventy years^28,29^. The *Danish National Patient Register* includes the full medical records of all individuals treated at Danish hospitals (inpatient department) since January 1, 1977, as well as in outpatient clinics since 1 January 1994 (or occasionally since 1995)^30^. The register was updated in 2002 to also include individuals treated in hospitals outside of Denmark and treatments not covered under the Danish health insurance agreement at private healthcare facilities. Finally, the *Danish Psychiatric Central Research Register* contains data on admissions to psychiatric inpatient facilities up to and including 1994. Following 1994, the register was extended to include outpatient contacts to psychiatric departments^31^. As of April 2017, the civil register contained 9,851,330 individuals, the national patient registers 8,065,597 individuals, and psychiatric register 1,005,068 individuals. All individuals were born between January 1, 1858, and April 21, 2017. All registers contained a unique personal identification number given to all individuals living in Denmark, therefore allowing for accurate linking across the different registers.

### Swedish nationwide registers

The *Swedish Total Population Register (TPR)*, started from 1968 and continuously updated, holds information on all individuals who are residents in Sweden. It contains information on birth, death, name change, marital status, family relationships and migration within Sweden as well as to and from other countries^38^. *Multi-Generation Register (MGR)*^39^ is part of TPR and contains information on all residents in Sweden who were born in 1932 or later and alive in 1961 (“index persons”), together with their parents. Familial linkage (i.e., parental information) is available for more than 95% of individuals who died before 1968, about 60% of those died between 1968 and 1990, and more than 90% of those alive in 1991. *The Swedish Inpatient Register* was launched in 1964 (psychiatric diagnoses from 1973) but complete coverage was reached since 1987. It includes discharge diagnoses, dates of hospital admission and discharge, and has a coverage of at least 71% of all residents for somatic care discharge in 1982 and 86% of all psychiatric care in 1973. Since 2001, this register also covers outpatient^40^. The individually unique National Registration Number was used to link data from all the registers. All Swedish-born residents were followed for any cardiometabolic and mental disorders from birth until emigration or death during 1973-2016.

### Register phenotypes

We defined the six MDs, namely attention-deficit/hyperactivity disorder (ADHD), anorexia nervosa (AN), autism spectrum disorders (ASD), affective disorders (AFF), bipolar disorder (BD), and schizophrenia (SCZ), and 14 cardiometabolic disorders, using information from the Danish and Swedish Patient Register. Note, that AFF includes two main diagnosis, BD and major depressive disorder. Individuals with at least one hospital visit concerning these disorders (primary or secondary diagnosis) were considered cases with MD or CMD. Individuals diagnosed with SCZ, BD, or AFF before age 10 were removed from the analysis, as the validity of such diagnosis is considered clinically unreliable. ICD 8 codes were used until 1993 and ICD 10 codes were used since 1994 in Denmark; ICD 8 codes were used until 1986, ICD 9 codes were used during 1987-1996, and ICD 10 codes were used since 1997 in Sweden (Table S9).

### GWAS summary statistics

A total of 14 cardiometabolic GWAS summary statistics including stroke (subtypes)^41^, CAD ^42^, aneurysms^43^ and HF^44^ were obtained through multiple public repositories. GWAS summary statistics containing participants of the VA Million Veterans Program (e.g. T2D^45^, venous thromboembolism^46^, and peripheral artery disease^47^) were provided after approval was granted by the National Institute of Health (*project #26508*). GWAS summary statistics for ADHD^48^, AN^49^, ASD^50^, BD^51^, and MDD^52^ excluding iPSYCH participants (except SCZ^53^ which does not contain iPSYCH samples) were kindly provided through their respective PGC consortium. iPSYCH only GWAS summary statistics for MDs^21^ were downloaded from internal iPSYCH servers and are available on request. The full list of all cardiometabolic- and mental disorder GWAS summary statistics used is shown in Table S6.

### GWAS summary statistics cleaning

All GWAS summary statistics were uniformly cleaned using internal software^54^. First, for each GWAS summary statistics we inferred the genome build by mapping SNPs to dbSNP build 151 using GRCh38, GRCHh35, GRCh36, and GRCh37 genomic coordinates. The version with highest number of mapped SNPs was inferred as the build of the original GWAS. Next, a second mapping step uses the inferred build to simultaneously map and liftover the position and chromosome coordinate to the GRCh37 version of dbSNP, which adds information about reference and alternative alleles. RSids were used when chromosome and base pair information were not available. The reference allele of dbSNP corresponds to the reference allele of the reference genome. The allele directions were flipped making the effect allele the reference allele. Effect scores (e.g., beta coefficients, odds ratios, and z-scores) were adjusted accordingly. Finally, multi-allelic, allele mismatched, and strand ambiguous SNPs alongside SNPs with duplicated positions, missing test statistics, and indels were removed^54^.

### LD-Score Regression

SNP based heritability (h^2^_SNP_) and genetic correlations (rg SNP) between all cleaned MD and CMD GWAS summary statistics were estimated using linkage-disequilibrium score regression (LDSC)^55,56^ using authors’ protocols.

### Cumulative Incidences

We estimated the cumulative incidence of all MDs and CMDs, which can be interpreted as the number of cases happening before a specific age. The cumulative incidences were estimated for the general population, individuals with one or more full siblings diagnosed with the same disorder, and individuals with one or more parents diagnosed with the cross-disorder (e.g., the cumulative incidence of ADHD for individuals with at least one parent diagnosed with type-2 diabetes). We expected the distribution of individuals into these 3 categories to be associated with birth year. Thus, to control for substantial changes over time in the underlying incidence, diagnoses (e.g., shifting of ICD systems), data availability, and registration (e.g., use of inpatient diagnoses up to 1995/2000 and in- and out-patient diagnoses subsequently), all cumulative incidences were estimated stratifying on the year of birth using the Nelson-Aalen estimator, which can utilize censored, competing risks, and incomplete data^18^. Next, we estimated the additive heritability (h^2^) and genetic correlation (rg) under the liability threshold model based on the cumulative incidence as a function of pedigree relatedness following procedures described by Wray and Gottesman^27,57,58^. In short, the liability threshold model assumes that disease liability underlying the disease status is normally distributed, Z∼N(0,1), and individuals with the disorder must therefore have surpassed a liability threshold^59,60^. Given the normal distribution theory the liability threshold of a given disorder can be estimated from the population that are affected in their lifetime (lifetime risk). All analyses were done in R v4.2.1 using the cmprsk v2.2 package.

### Heritability (h^2^)

Using the full available register data (no restriction of birth year), the heritability of liability of disorders was calculated by deriving the general population-(e.g., risk of ADHD in the population) and full-sibling familial risk (e.g., risk of ADHD when having a full-sibling with ADHD) cumulative incidences for individuals born in the same calendar year (e.g., 1965, 1966, till 2016). Here, we use the cumulative incidence (general population and full-sibling risk) at the last observed time point as estimates of the proportion of the population born in the same calendar year that are affected in their lifetime resulting in estimates of heritability (Equations A1 and A2) for individuals born in the same calendar year (ℎ^2^*_year of birth_*).

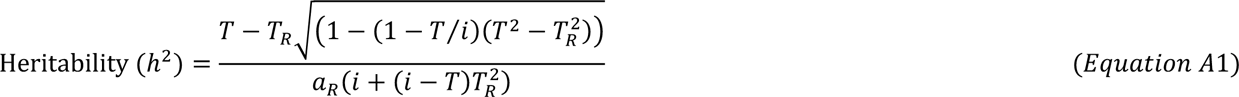

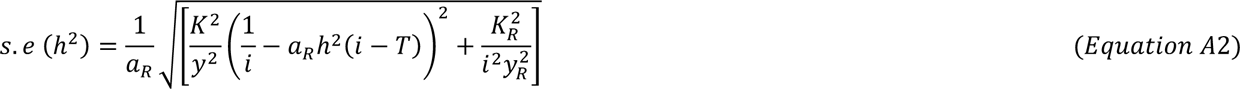

*Where T=Liability threshold of the disease in the general population, T_R_=Liability threshold of the disease based on affected family members, i=mean liability of disease in the population calculated as i=y/K; where K is the lifetime probability of disease in the population and y the height of the normal curve at threshold T, a_R_ =additive genetic relationship between relatives,* K_R_ *= the lifetime probability of disease in individuals with affected family members. Note that all estimates derived for individuals’ born in the same calendar year*.

### Genetic correlation (rg)

In contrast to the h2 estimation, for the genetic correlation we restricted the birth window to individuals born between 1981 and 2005, using medical records up to 2012. The rg between disorders was calculated by deriving: the general population risk for both disorders (e.g., ADHD and T2D) and parent-offspring cross disorder familial risk (e.g., risk of ADHD when having a parent with T2D) cumulative incidences for individuals born in the same calendar year (e.g., 1981,1982 till 2005). In line with the h^2^ estimation we used the cumulative incidence at the last observed time point for each birth year for all three cumulative incidence functions (general population risk and cross disorder familial risk). Using the h^2^ of both disorders previously obtained we derived estimates of genetic correlations (Equations B1 and B2) per year of birth (*r*_*g,year of birth*_).

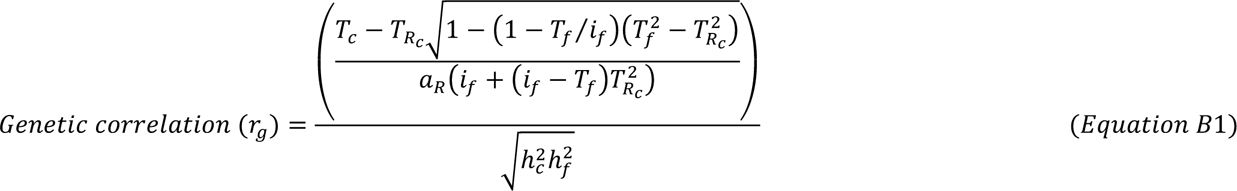

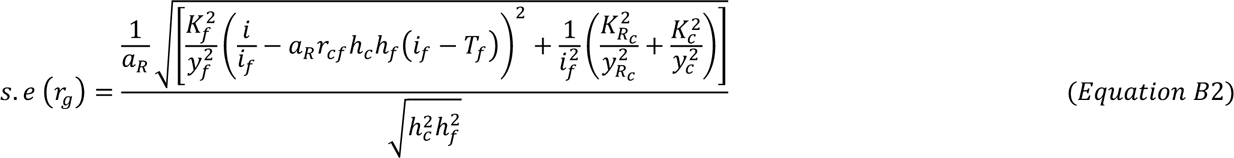

Where T_c_ and T_f_ =*Liability threshold of disease c and f in the general population, T*_*R_c_*_ *= Liability threshold of disease c in individuals with relatives with disease f, i_f_=mean liability of disease f in the population, a_R_ =additive genetic relationship between relatives, h*^2^*_c_* and *h*^2^*_f_* = heritability of diseases c and f, K_f_ *is the lifetime probability of disease f in the general population. Note that all estimates derived for individuals’ born in the same calendar year*.

### Random effect inverse variance weighted model (IVWrandom)

We obtain overall h^2^ and rg estimates by weighing the individual *h*^2^_*year of birth*_ and *r*_*g, year of birth*_ by the inverse of their sampling variance (Equations C1 and C2) using a random-effects model.

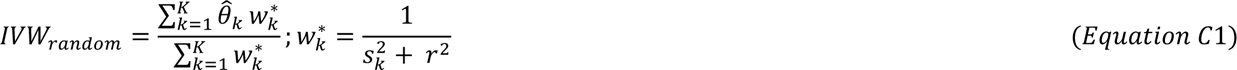

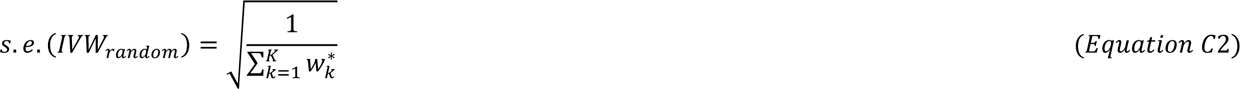

Where K = numbers of estimates, *s*^2^*_k_* = *variance of estimate* k, r^2^ = the variance of the distribution of true effect sizes, *θ^^^_k_* = *point estimate* k, and *w*^*^*_k_* = random-effects weight.

### Quantification of genetic and non-genetic factors

Under a bivariate liability threshold model, the phenotypic correlation (rP) between two traits can be broken down to its (additive) genetic- and non-genetic factors. This allows us to quantify and understand the contribution of the estimated genetic correlation and heritability to the level of comorbidity between MDs and CMDs, i.e., hazard ratios) reported by Momen et al^1^, which uses the same Danish register data.

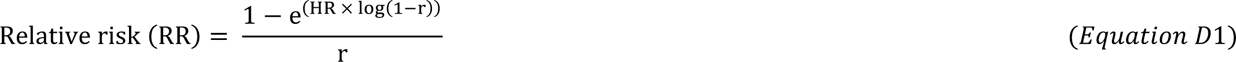

*Where HR =hazard ratio reported by Moment et al, 2020, and r = rate of the disorder in the reference group derived by weighting the individual estimates (1981-2005 using medical records up to 2012) by the inverse of their sampling variances*.

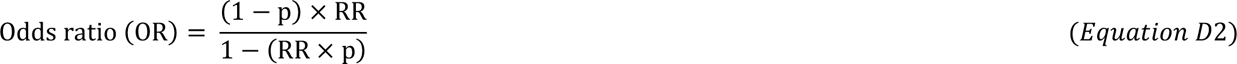

*Where p = incidence of the disorder in the nonexposed group (here p=r) and RR = the calculated relative risk*.

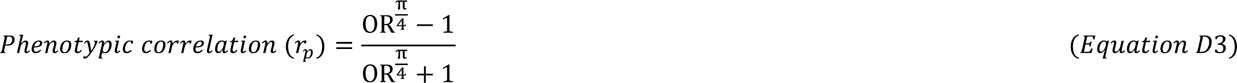

*Where OR = odds ratio estimated as a function of relative risk*.

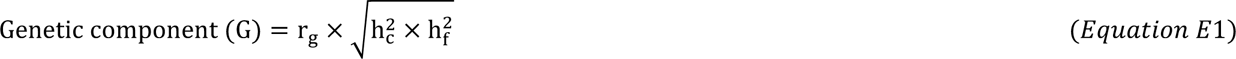

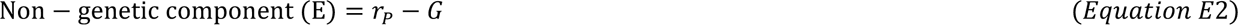

*Where r_p_ = tetrachoric correlation derived from the HR, r_g_ = the genetic correlation estimates between disorder c and f, h^2^ the heritability estimates for c and f. 95% CIs for G and E were derived using both the upper and lower 95% CIs of r_g_ and r_p_. Note to estimate G_SNP_ and E_SNP_ replace r_g_ and h^2^ estimates by SNP based estimates r_g SNP_ and h^2^_SNP_*.

## Supplementary Figures

**Supplementary figure S1:**
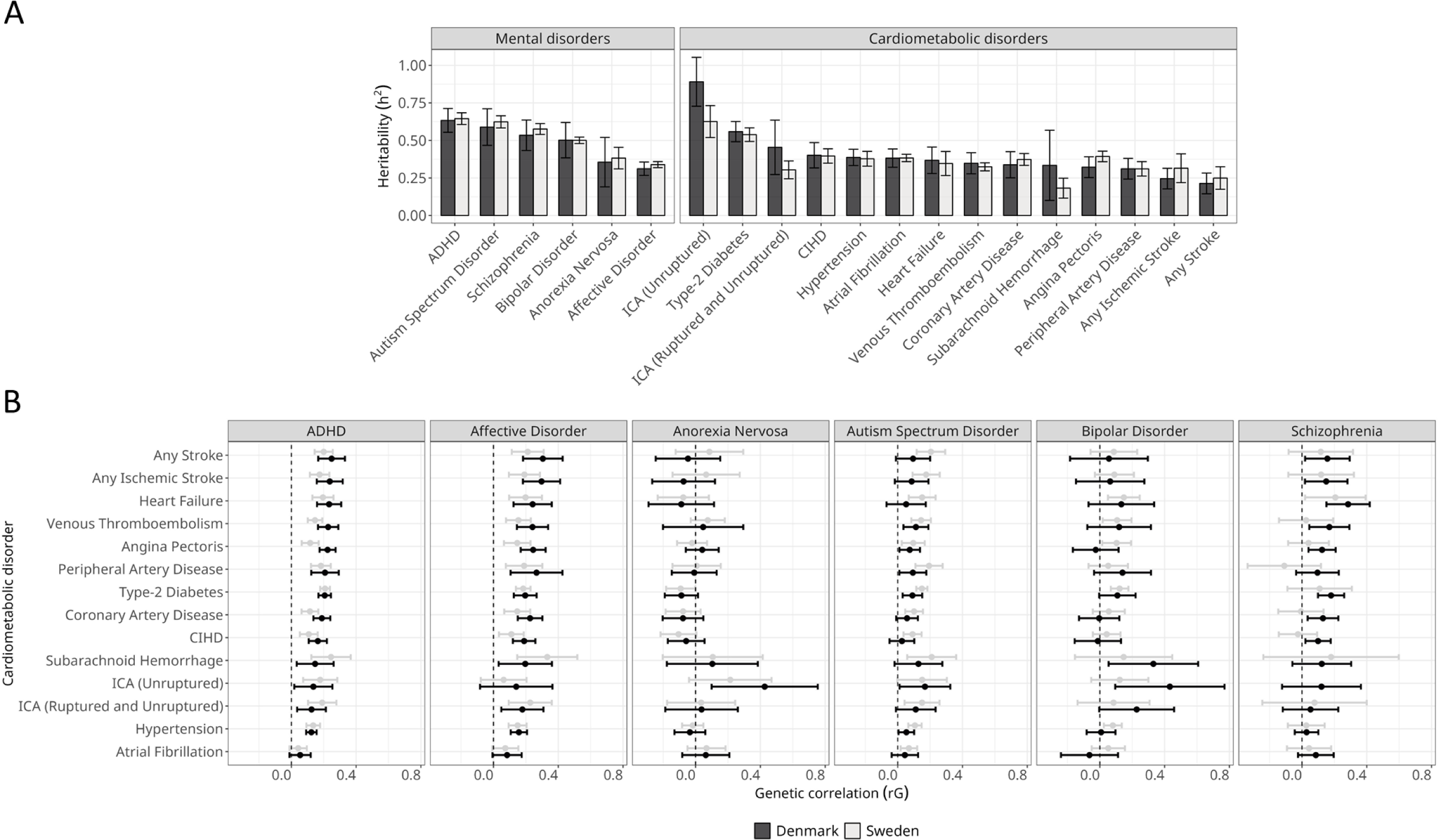
Comparison of Danish and Swedish register-based estimates and 95% confidence intervals for A.) heritability (h^2^) B.) genetic correlations (rg). *ADHD = attention deficit/hyperactivity disorder, ICA = intracranial aneurysm, CIHD = chronic ischemic heart disease*

**Supplementary figure S2:**
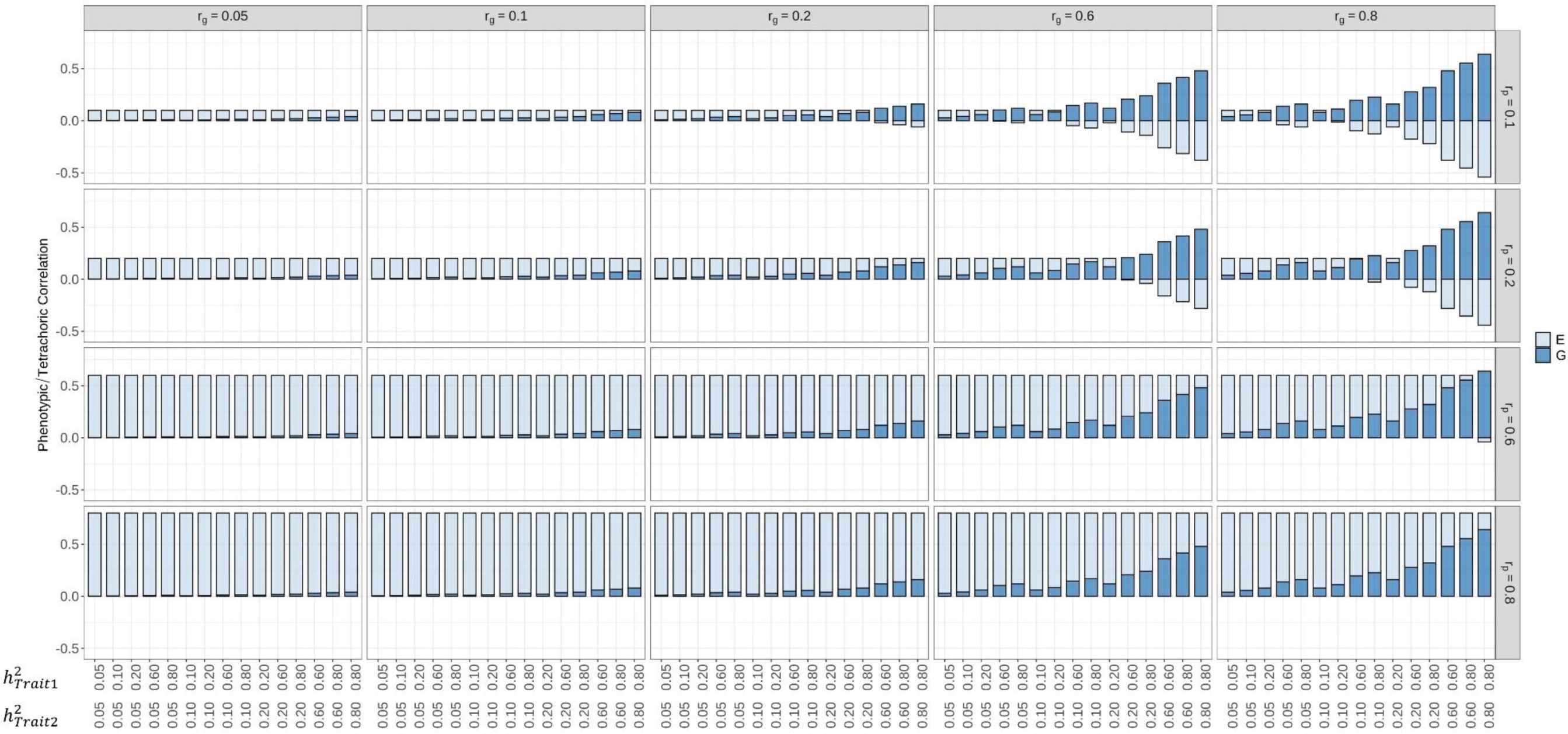
The effect of heritability (h^2^) and genetic correlation (rg) size on the magnitude of the genetic (G) and non-genetic (E) contribution to the phenotypic correlation (rp) between two traits. *Phenotypic correlations (rp) were derived using simulated hazard ratios (HZ) and prevalence’s (Pr) of: rp = 0.1: HZ=1.28, Pr=0.05; rp=0.2: HZ=1.61, Pr=0.12; rp=0.6: HZ=4, Pr=0.2; rp=0.8: HZ=4, Pr=0.5*.

**Supplementary figure S3:**
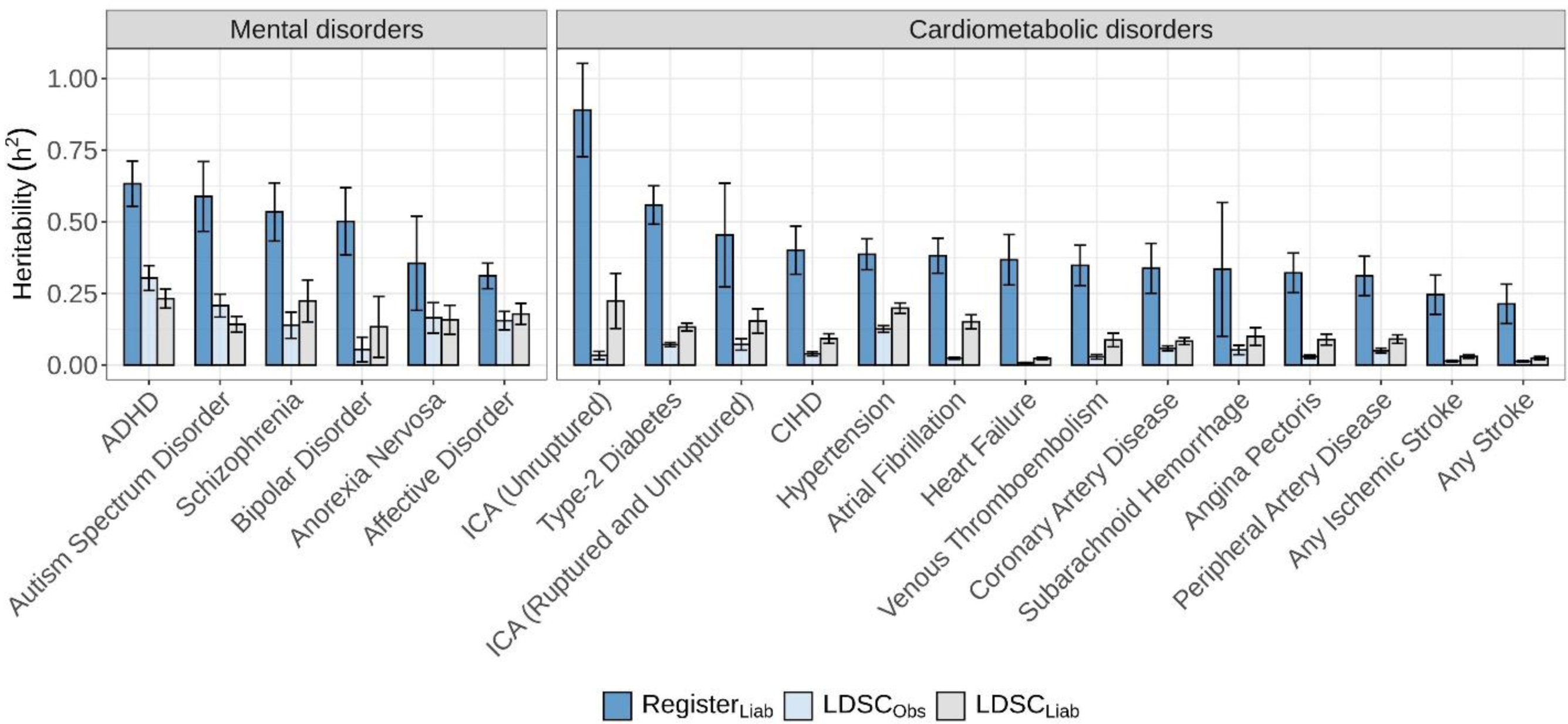
Narrow sense heritability (h^2^) estimates and 95% confidence intervals of mental- and cardiometabolic disorders using the Danish registers (RegisterLiab) and LD-score regression on the observed (LDSCObs) and liability scale (LDSCLiab). *ADHD = attention deficit/hyperactivity disorder, ICA = intracranial aneurysm, CIHD = chronic ischemic heart disease*

**Supplementary figure S4:**
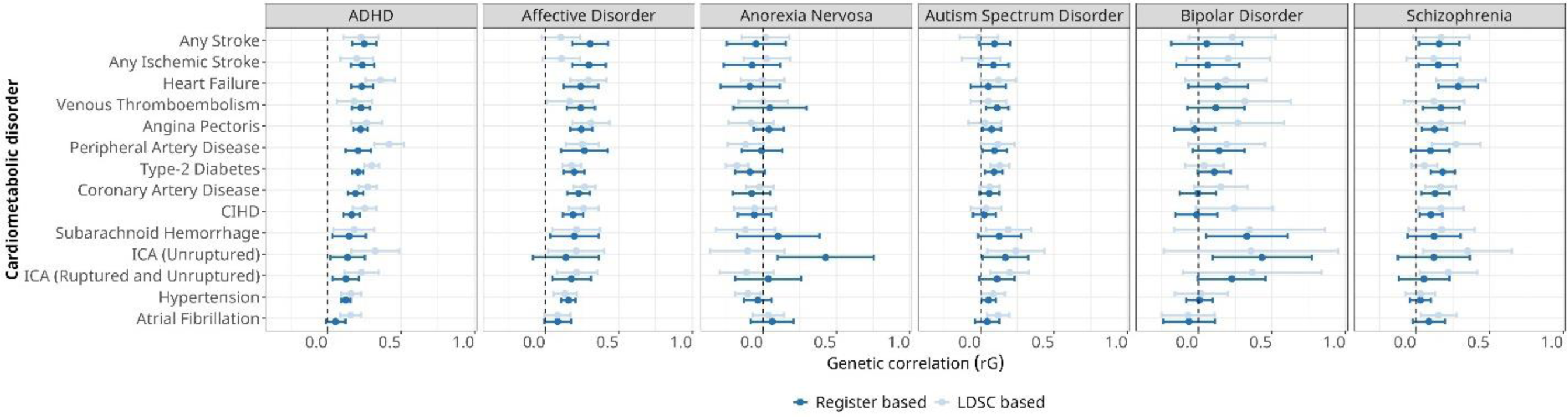
Comparison of the genetic correlations and 95% confidence intervals between mental- and cardiometabolic disorders calculated using register data (rg) and LDSC (rg SNP). *ADHD = attention deficit/hyperactivity disorder, ICA = intracranial aneurysm, CIHD = chronic ischemic heart disease*

**Supplementary figure S5:**
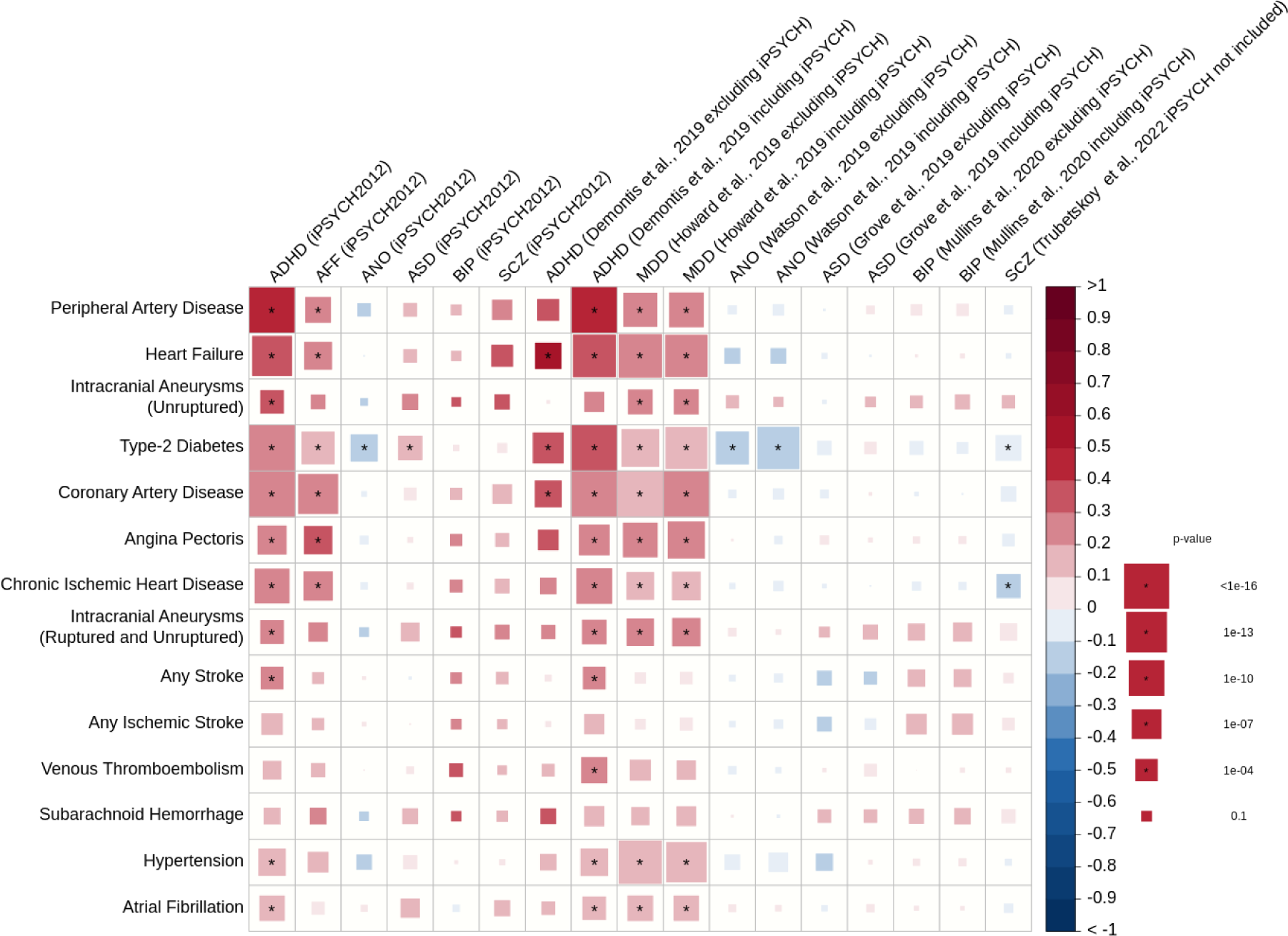
LDSC genetic correlations between six mental- and 14 cardiometabolic disorders. Genetic correlations (rg SNP) with standard error, and p-values. *We repeated the analysis three times: using only iPSYCH samples and using the Psychiatric Genomics Consortium (PGC) meta-analysis including and excluding iPSYCH samples. Trubetskoy et al, 2022 did not contain iPSYCH samples. No PGC-AFF disorder GWAS meta-analysis exists therefore we used MDD as the next closest disorder*.

**Supplementary figure S6:**
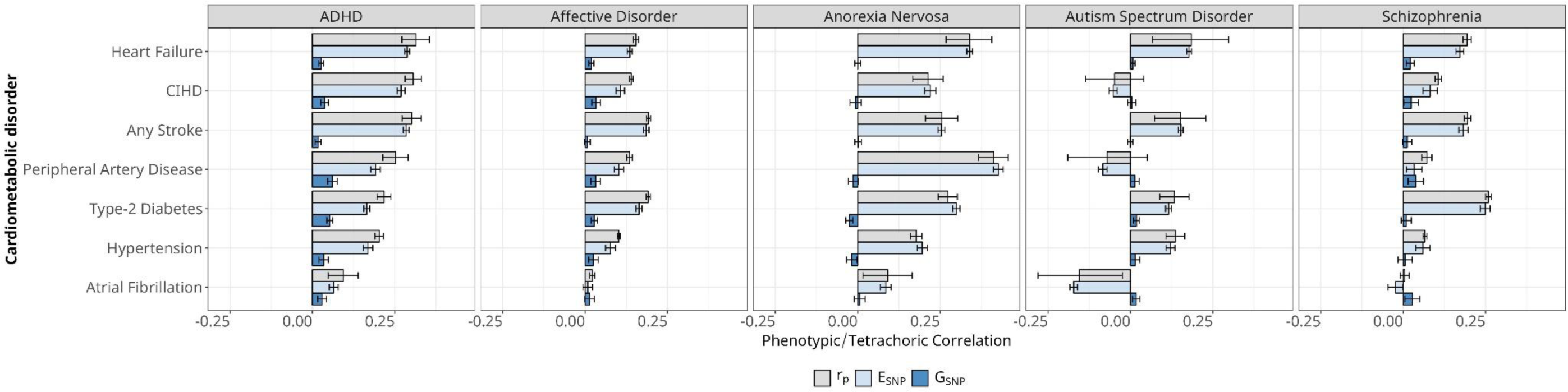
Quantification of the contribution of genetic (GSNP) and the non-genetic (ESNP) factors to the observed phenotypic correlation. (rp) **between mental- and cardiometabolic disorders using LDSC based genetic correlations (rg SNP) and heritability (h^2^SNP) estimates.** *Estimates of rp were selected from Momen et al.^1^*.

## Notes

### Funding Statement

This research was supported by: the European Unions Horizon 2020 Research and Innovation Programme: the "predicting comorbid cardiovascular disease in individuals with mental disorder by decoding disease mechanisms" project (CoMorMent, grant number 847776, to JM, JB, AB, UAV, DHM, YL, TMW, OAA and FF); the Danish National Research Foundation (grant number DNRF148); the US National Institutes of Health study on extreme MDD (R01 MH123724, to JM, JRS, YL, and AB); the Sir Henry Wellcome Postdoctoral Fellowship (Reference 213674/Z/18/Z, to DMH); the Research Council of Norway (RCN grants 324499, 324252, 223273, to OAA); the Stiftelsen Kristian Gerhard Jebsen (grants SKGJ-MED-008 and SKGJ-MED-021, to OAA); the Laureate Grant Award from the Novo Nordisk Foundation (Grant No: NNF22OC0071010, to NM); European Research Council Consolidator grant (StressGene, Grant nr. 726413 to UAV), and Icelandic Research fund (to UAV). The iPSYCH Initiative is funded by the Lundbeck Foundation (Grant Nos. R268-2016-3925, R102-A9118, and R155-2014-1724), the Mental Health Services Capital Region of Denmark, University of Copenhagen, Aarhus University, and the University Hospital in Aarhus. Genotyping of iPSYCH samples was supported by grants from the Lundbeck Foundation, the Stanley Foundation, the Simons Foundation (Grant No. SFARI 311789), and National Institutes of Mental Health (Grant No. 5U01MH094432-02). The iPSYCH Initiative uses the Danish National Biobank resource that is supported by the Novo Nordisk Foundation.

